# Gestational diabetes mellitus in women born small or premature: Systematic review and meta-analysis

**DOI:** 10.1101/2021.06.29.21259746

**Authors:** Yasushi Tsujimoto, Yuki Kataoka, Masahiro Banno, Shunsuke Taito, Masayo Kokubo, Yuko Masuzawa, Yoshiko Yamamoto

## Abstract

**Background:** Women born preterm or with low birthweight (LBW) have an increased future risk of gestational diabetes mellitus (GDM) during pregnancy; however, a quantitative summary of evidence is lacking.

In this study, we aimed to investigate whether being born preterm, or with LBW or small for gestational age (SGA) are associated with GDM risk.

**Methods:** We searched the MEDLINE, Embase, and CINAHL databases and study registries, including ClinicalTrials.gov and ICTRP, from launch until 29 October 2020 for observational studies examining the association between birth weight or gestational age and GDM were eligible. We pooled the odds ratios and 95% confidence intervals using the DerSimonian and Laird random-effects model.

**Results:** Eighteen studies were included (N = 827,382). The meta-analysis showed that being born preterm, with LBW or SGA was associated with increased risk of GDM (pooled odds ratio = 1.84; 95% confidence interval: 1.54 to 2.20; I^2^ = 78.3%; τ^2^ = 0.07). Given a GDM prevalence of 2.0%, 10%, and 20%, the absolute risk differences were 1.6%, 7.0%, and 11.5%, respectively. The certainty of evidence was low due to serious concerns of risk of bias and publication bias.

**Conclusion:** Women born prematurely, with LBW or SGA status, may be at increased risk for GDM. However, whether this should be considered in clinical decision-making depends on the prevalence of GDM.

**What is already known on this subject?:** Approximately 10–15% of infants are born small or premature worldwide, and these children are at high risk of diseases, such as type 1 and 2 diabetes mellitus, hypertension, obesity, and kidney disease, in adulthood. A narrative review reported in 2007 that women born with LBW are at risk of gestational diabetes mellitus, but included a small number of studies. Several subsequent studies have been published since then, but there is no quantitative summary of the relevant evidence to date.

**What this study adds?:** This is the first systematic review and meta-analysis of observational studies that provides a comprehensive summary of evidence on the association between birth size or premature birth and future GDM risk including previously unpublished data and a large sample size. LBW, preterm birth, and SGA status may be prognostic factors for GDM.

## INTRODUCTION

Gestational diabetes mellitus (GDM) is a common pregnancy complication, with prevalence estimates being 1–36%, depending on the population studied and diagnostic criteria employed ^1^. GDM is defined as preconceptionally unconfirmed glucose intolerance identified in the second or third trimester of pregnancy ^2^. Adverse perinatal outcomes associated with uncontrolled diabetes in pregnancy include spontaneous abortion, foetal anomalies, preeclampsia, stillbirth, macrosomia, neonatal hypoglycaemia, and neonatal hyperbilirubinemia, among others ^3^. Women with a history of GDM are at a higher risk of type 2 diabetes than their counterparts ^4, 5^.

Low birth weight (LBW) and preterm birth are the leading causes of neonatal death and childhood-onset morbidity ^6^. Approximately 10–15% of infants are born small or premature worldwide ^6, 7^. Children who survive are at a higher risk of diseases, such as type 1 and 2 diabetes mellitus, hypertension, obesity, and kidney disease, in adulthood ^8^. The exact mechanism underlying these risks remains unclear; the Barker hypothesis proposes that pregnancy may activate biological vulnerability in utero ^9^.

A narrative review reported in 2007 that women born with LBW are at risk of GDM ^10^, but included a small number of studies, and additional research has been published subsequently ^11, 12^. There is no quantitative summary of the relevant evidence to date. We performed the first systematic review and meta-analysis of observational studies examining the association between preterm birth, with LBW, or with SGA status and the future risk of GDM.

## METHODS

We followed the Meta-analysis of Observational Studies in Epidemiology (MOOSE) guidelines (Supplementary Table S1) in the reporting of this study; the study methodology adhered to the Cochrane Handbook ^13, 14^. Evidence certainty assessment was based on the Grading of Recommendations, Assessment, Development and Evaluation (GRADE) criteria for prognostic factors ^15^. The protocol was prospectively registered with PROSPERO (CRD42020142004).

### Searches

We searched databases such as MEDLINE, Embase, and CINAHL and study registries including ClinicalTrials.gov and ICTRP from launch until 29 October 2020. Qualified authors (YT and YK) developed the search strategy (Supplementary Table S2). No language or publication status restrictions were imposed. Reference lists of shortlisted studies were searched manually for additional potentially eligible titles.

### Study selection

Studies were eligible for inclusion if they were observational cohort or case-control studies. Case reports or series were excluded from the present review. We included studies that involved pregnant women regardless of study setting. The exposures of interest were the infancy parameters of presently pregnant women and were defined as follows: LBW, birth weight <2500 g ^7^; small for gestational age (SGA), birth weight <10th percentile for the given gestational age, stratified by sex, using the average weight of gestational age ^16^; and preterm birth, gestational age of <37 weeks ^17^. When data on both birth weight and gestational age were reported, we extracted data on birth weight in preference. The comparator group comprised women who were not born small or born at full term.

The outcome of interest was GDM, as defined by the International Association of Diabetes Pregnancy Study Groups (IADPSG), World Health Organization (WHO), American Diabetes Association or Endocrine Society, or International Classification of Diseases 11th revision (ICD-11) or earlier ^18–22^. If studies used other definitions, they were included in the present review; however, we removed them to assess the robustness of the pooled estimates. For studies that reported LBW, preterm birth, or SGA as a risk factor in pregnant women without reporting the association with GDM, we contacted study authors to acquire estimates of such associations, where available. These additional estimates were included in the present analysis, provided they were measures of an association between at least one of the exposure factors and the outcome of interest.

Two investigators independently screened article titles and abstracts to shortlist relevant studies; subsequently, the same sets of authors assessed the full text for study eligibility. In cases where data were incomplete and precluded study eligibility assessment, we contacted study authors with requests for clarification. Multiple publications were assessed together; the record with the most complete data was included in the present review.

### Data extraction and quality assessment

Two investigators independently extracted data from all included studies, using a pilot-tested, uniform data extraction sheet. Any discrepancies between reviewers were resolved through consensus between two reviewers or arbitration by a third reviewer, as required. For studies that compared three or more exposure groups, we contacted study authors to obtain data comparing two groups of interest. In cases where this approach was unsuitable, we extracted the relevant data, as reported, and performed subgroup comparisons between the two groups subdivided by specific thresholds (i.e., birth weight 2500 g, <10th percentile, and gestational age 37 weeks for LBW, SGA, and preterm birth, respectively), as this approach may have resulted in conservative effect estimates. The same authors who performed data extraction also independently assessed the risk of bias in each study, using the Quality In Prognosis Studies (QUIPS) tool ^23^. We prospectively identified the following candidate confounders: age, obesity, smoking status, socioeconomic status, diabetes mellitus before the index gestation, and family history of diabetes ^24, 25^.

### Data synthesis and analysis

We obtained pooled and adjusted ORs with 95% CI estimates of GDM for the exposure and control groups using the DerSimonian and Laird random-effects method. We calculated the absolute risk difference for GDM between the exposure and control groups in low-(control group: GDM risk was assumed to be 2.0%), moderate- (10%), and high- (20%) prevalence groups, using the pooled odds ratios (OR) and 95% confidence intervals (CIs). This assumption was made based on a previous report and our clinical expertise ^26^.

Publication bias was assessed qualitatively by visual inspection of the funnel plot and quantitatively by Egger’s test ^27^. Where asymmetry was observed in the funnel plot, we investigated the likely source of this asymmetry using the contour-enhanced funnel plot. We evaluated between-study heterogeneity visually, using forest plots, and quantitatively, using *I*^2^ and τ^2^ statistics. We used the Cochrane chi-square test to calculate *I*^2^ and τ^2^ statistics. We performed a pre-specified subgroup analysis based on types of exposure (preterm birth, LBW, or SGA). In pre-specified sensitivity analyses, we used crude ORs instead of adjusted ORs and excluded studies using non-standard definitions of GDM. Some studies assessed the risk of GDM among women born with a weight >4000 g (macrosomia); these studies were excluded from post-hoc sensitivity analysis, as a previous review has shown a U-shaped association between mother’s birth weight and GDM risk ^10^.

All analyses were performed using STATA 14.2 (StataCorp LP, Texas) and RevMan 5.4 (Cochrane Collaboration, UK). Two-sided *p-*values <0.05 were considered indicative of statistical significance.

## RESULTS

Figure 1 presents the flow of studies through the present review selection process. After screening 15,281 records, 59 records representing 44 studies were assessed for eligibility based on the full text. Finally, 18 studies including 827,382 participants were included in the qualitative synthesis; 15 studies including 825,622 participants were included in quantitative synthesis. Supplementary Table S3 lists all excluded studies with reasons for exclusion. We did not find any ongoing or unpublished studies by searching study registries. By contacting authors, we obtained unpublished data from two studies ^12, 28^.

**Figure 1.**
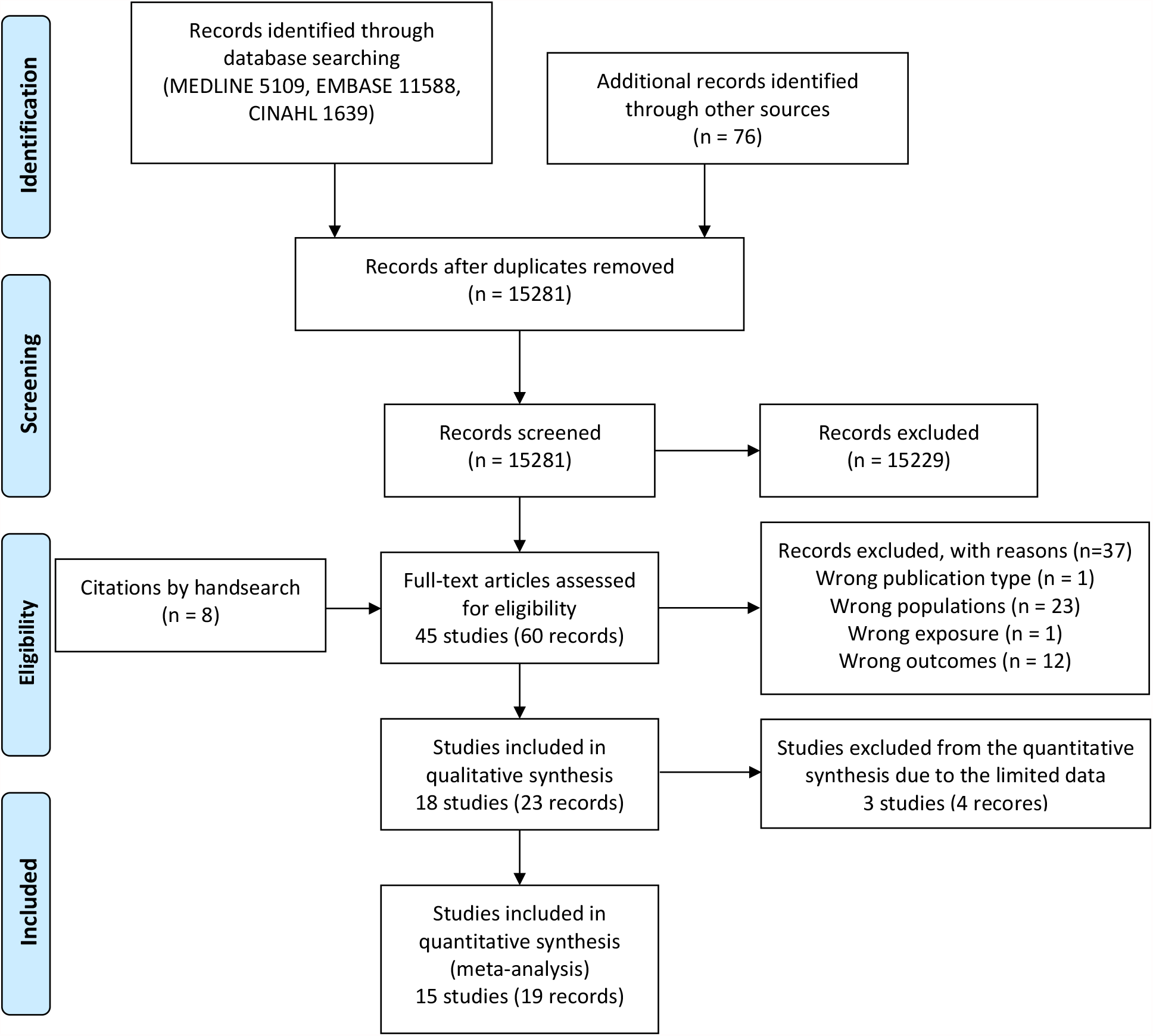
PRISMA flow diagram of study eligibility. Duplicate studies are displayed as a single study.

Supplementary Table S4 shows the detail characteristics of the included studies. Nine studies (810,197 participants) used population-based samples, 2 (6,915 participants) were multicentre studies, 6 (9,439 participants) were single-centre studies, and 1 (831 participants) did not specify the study setting. Supplementary Table S5 shows the details of the inclusion and exclusion criteria of the included studies. All studies were conducted in high-income countries, mostly between the late 1990s and early 2010s. The studies included participants of non-Hispanic White, Hispanic, African, Asian, or Indian descent. Two studies (28,722 participants) only included women about to deliver their first child. Two studies (140,714 participants) compared pregnant women born preterm and at full term, 9 (216,439 participants) compared women born with and without LBW, and 4 (468,469 participants) compared women born with and without SGA status. The remaining 3 studies (1,760 participants) only compared the mean birth weight of women with or without GDM. Figure S1 presents a summary of study quality assessment using the QUIPS tool ^23^. The overall quality of the included studies was moderate to low, mainly due to uncontrolled confounders.

### Prematurity and size at birth and the risk of gestational diabetes mellitus

The median GDM rate in the control groups of the included cohort studies was 2.9% (range: 0.5% to 22%). Figure 2 presents a forest plot summarising the studies that assessed the association between preterm birth or size at birth with GDM. Premature birth, LBW, and SGA status were associated with a higher GDM risk (pooled OR, 1.84; 95% CI: 1.54 to 2.20; I^2^ = 78.3%; τ^2^ = 0.07). Supplementary Table S5 summarises the absolute risk difference in pregnant women born with LBW, SGA status, or born preterm in the low- (2.0% risk of GDM in the control group), medium- (10%), and high- (20%) GDM prevalence groups. The absolute risk increases were 1.6% (95% CI: 1.0 to 2.1%), 7.0% (95% CI: 4.6 to 9.6%), and 11.5% (95% CI: 7.8 to 15.5%) in low-, moderate-, and high-prevalence settings, respectively. The certainty of evidence was low due to serious concerns of risk of bias and publication bias.

**Figure 2.**
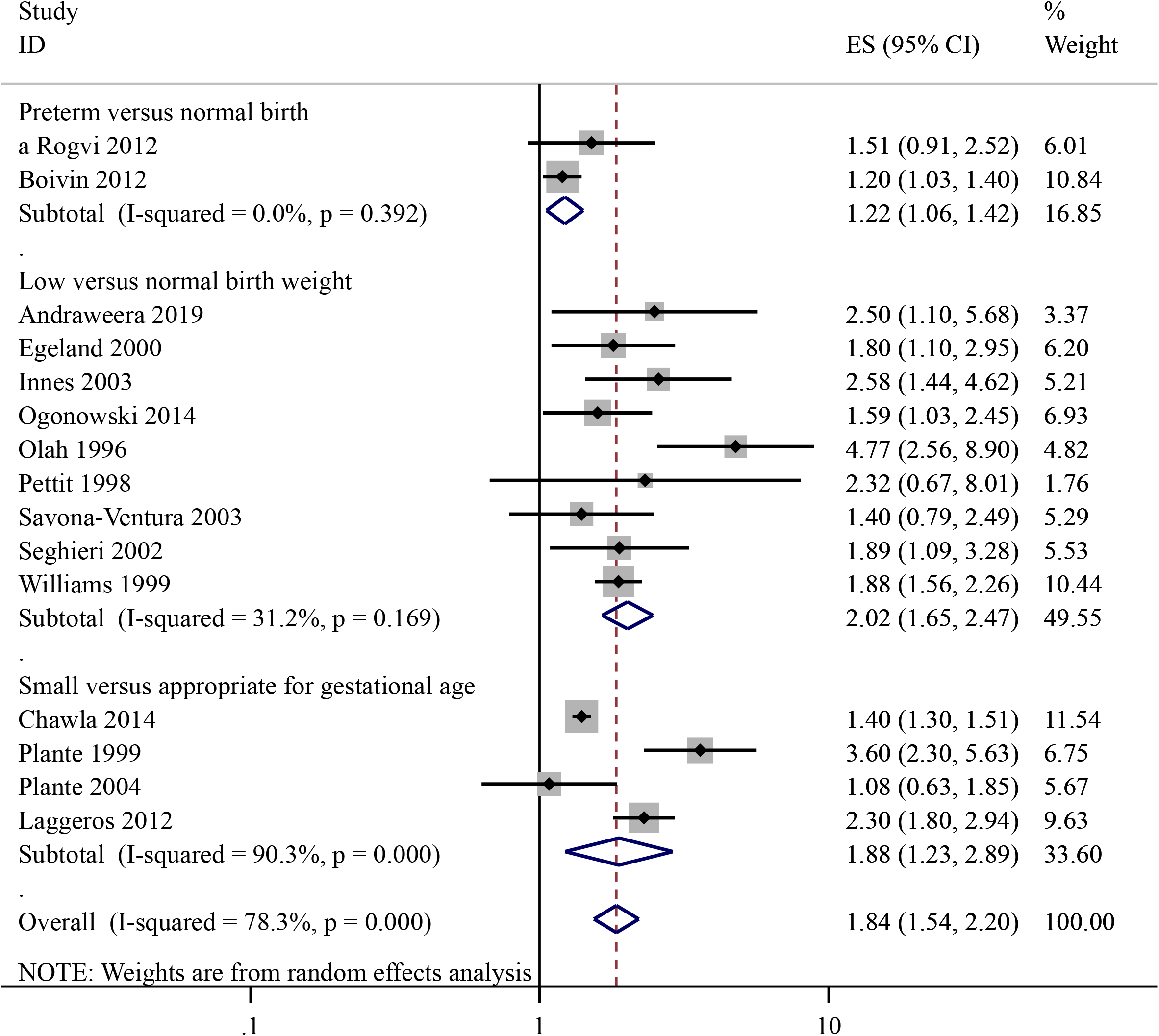
Risk of gestational diabetes among women born preterm, with low birth weight, or small-for-gestational-age status. Effect size (ES, represented as adjusted odds ratios); CI, confidence interval. ES was determined using the random-effects model weighted by the inverse of the variance estimate. Squares represent ES, with marker size reflecting the statistical weight of the study, obtained using random-effects meta-analysis; horizontal lines represent 95% CIs; diamonds represent the subgroup and overall odds ratios and 95% CIs for gestational diabetes.

Figure 3 presents study estimates in a funnel plot. The plot appeared asymmetrical, and Egger’s test for funnel plot asymmetry was statistically significant (*p*-value = 0.030). Supplementary Figure S2 shows the contour-enhanced funnel plot, which suggests the existence of some missing studies on the left-hand side of the plot; these studies would have yielded statistically non-significant findings.

**Figure 3.**
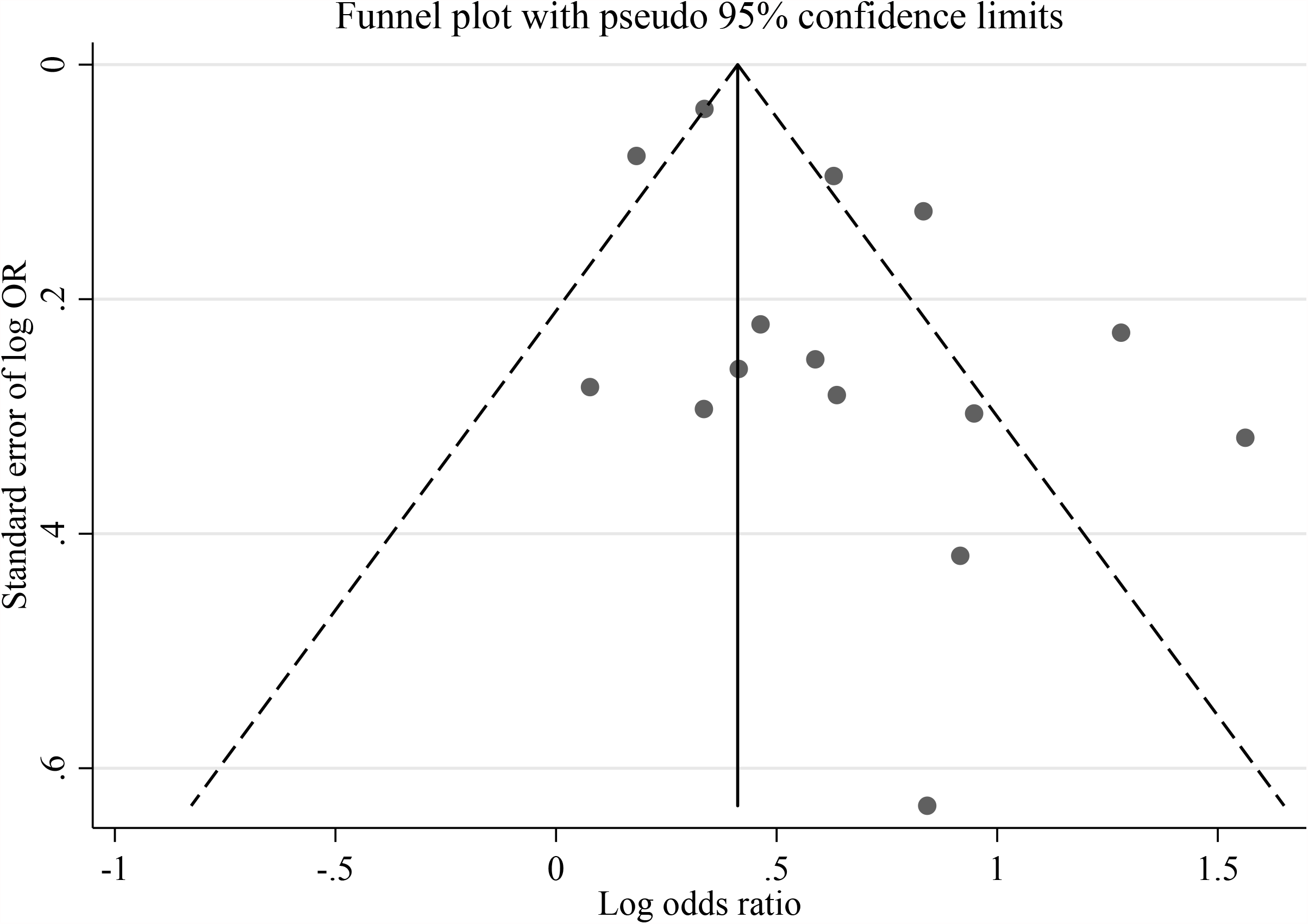
Funnel plot for the evaluation of publication bias. The solid vertical line represents the summary estimate of the association between preterm birth, low birth weight, and small for gestational age status and gestational diabetes (using random-effects meta-analysis). A significant publication bias was detected (*p*□=□0.030 for Egger’s test). The funnel plot shows asymmetry, which indicates publication bias.

Data on the birth weight of mothers with or without GDM, obtained from three studies excluded from the meta-analysis, are presented in Supplementary Table S5. These studies consistently reported that mothers with GDM were born with lower birth weights than those without GDM.

### Subgroup and sensitivity analyses

There was substantial between-study heterogeneity (*I*^2^ = 78.3%). Figure 2 presents the results of subgroup analyses for the types of exposure (LBW, preterm birth, or SGA). Although all types of exposure were associated with GDM, there was significant heterogeneity due to the type of exposure (*p* for interaction = 0.004). The results of additional sensitivity analyses are presented in Supplementary Figure S3, Figure S4, and Figure S5.

## DISCUSSION

### Main Findings

We found that women born small or premature may have future risk of GDM. However, the evidence certainty was low, and the presented findings may be overestimated, as we observed some evidence of publication bias. These findings were approximately consistent across the subgroups, including different populations, exposures, and studies of varied methodological quality; these findings were robust in sensitivity analyses.

Our finding that the mother’s size at birth or premature birth may affect GDM risk was consistent with that of a previous narrative review ^10^. The strength of this association was similar to that observed in women with a family history of diabetes mellitus, an established risk factor for GDM ^29^. However, the importance of the risk factor in clinical decision depends on the absolute risk difference. Our findings suggested that careful review of the mother’s birth status may indicate her risk of GDM and guide pregnancy management in moderate to high prevalence settings. The mother’s preterm birth status and size at birth are not currently considered risk factors for GDM in any of the major guidelines or risk models ^30, 31^. Our findings may help further refine these guidelines and models or to develop new ones.

The certainty of evidence for the association between premature birth or SGA status and GDM was low due to the high risk of publication bias, as shown by funnel-plot analysis. The contour-enhanced funnel plot suggests that studies with non-significant findings may not have been published. Although we did not identify any ongoing or unpublished studies, this did not eliminate the risk of publication bias, as observational studies are less likely to be registered than clinical trials ^32^. Thus, the reported estimates may be overestimates. The studies included in this review tended not to adjust for confounders, such as smoking, obesity, socioeconomic status, and family history of diabetes. Future studies should adjust for these factors.

The main result of this review was subject to substantial between-study heterogeneity, as shown by the *I*^2^ statistic ^13^. This heterogeneity may be due to the different types of exposure (LBW, SGA, or preterm birth) considered in this study. However, as all exposure types were associated with increased GDM risk, the high *I*^2^ statistic may be due to the large number of participants and narrow CIs of the primary studies ^33^. Given these findings, we did not assign a low rating to the inconsistency domain of the GRADE criteria ^15^.

The underlying mechanism of the association between preterm birth or SGA status and subsequent GDM may be gestational malnutrition due to maternal malnutrition or placental insufficiency ^34^. Findings from animal studies have suggested that malnutrition in utero is associated with reduced β-cell counts, pancreas weight, and pancreatic insulin content ^35^. According to the Barker foetal origin hypothesis, these foetal programming events may affect the future risk of disease ^9^. A review of epidemiological studies has suggested that LBW and preterm birth are associated with the risk of type 2 diabetes in adulthood; a similar mechanism is possible for GDM ^36^.

### Strengths and Limitations

A key strength of this review is that it is the first to provide a comprehensive summary of evidence on the association between birth size or premature birth and future GDM risk. This study followed the methodological recommendations presented in the Cochrane Handbook, MOOSE guidelines, and GRADE criteria ^13–15^. Moreover, this study included previously unpublished data and a large sample size.

Nevertheless, this study has some limitations. First, the included studies were old and may not represent the current clinical practice. The definition of GDM proposed by the IADPSG in 2010 has resulted in an increase in GDM prevalence ^37^. For example, the prevalence of GDM in the United States increased from 4.6% in 2006 to 8.2% in 2016 ^38^. The median prevalence of GDM in the control groups of the included studies was 2.9%. However, empirical evidence suggests that relative effect measures are, on average, consistent across different settings; in the present study, we estimated absolute risk differences separately for low-, moderate-, and high-prevalence settings ^39^. Second, 5 of 15 studies divided birth size and preterm birth categories into three or more comparative groups, which could not be combined into two comparison groups of interest. This lack of data required methodological adjustments, as described previously. Lastly, this review only assessed certainty in estimates of association between prognostic factors and an outcome. Future studies are required to determine whether these factors can help risk-stratify pregnant women and improve the clinical management of GDM.

## Conclusions

LBW, preterm birth, and SGA status may be prognostic factors for GDM. Clinicians should consider the prevalence of GDM in their setting when considering maternal preterm birth or size at birth in clinical decision-making. Due to the high likelihood of publication bias, the true association between the exposures and outcome of interest may be weaker than that reported herein. Future studies based on up-to-date diagnostic criteria, examining the dose–response relationship between exposure severity and outcome, and comparing low-and middle-income countries, are required to improve the certainty of evidence.

## Data Availability

Data are available upon reasonable request.

## ACKNOWLEDGMENTS

We wish to thank Prof. Anne Monique Nuyt, Dr. Ariane Boivin, Dr. Prabha H. Andraweera, and Dr. Shalem Leemaqz for kindly providing additional study data that were not included in the original reports. We are also grateful to Dr. Kyosuke Kamijo and Dr. Ayako Shibata for contributing a clinical perspective. We would like to thank Editage (www.editage.com) for English language editing.

## CONTRIBUTION TO AUTHORSHIP

YT is the guarantor of the review. YT had full access to all of the data in the study and takes responsibility for the integrity of the data and the accuracy of the data analysis.

Study concept and design: YT, YK, MB, ST, MK, YY

Acquisition, analysis, or interpretation of data: All authors

Drafting of the manuscript: YT, YK, MB, ST

Critical revision of the manuscript for important intellectual content: MK, YM, YY

Statistical analysis: YT

Administrative, technical, or material support: YT, YK, MB, ST

Study supervision: MK, YM, YY

