## Supplementary for "Gestational diabetes mellitus in women born small or premature: Systematic review and meta-analysis"

Online Supplementary Material

**Supplementary Table S1. MOOSE checklist for the review**

**Supplementary Table S2. Search strategies**

**Supplementary Table S3. List of studies excluded from this review and reasons for exclusion**

**Supplementary Table S4. Detail characteristics of included studies**

**Supplementary Table S5. Details of inclusion and exclusion criteria for the original studies**

**Supplementary Table S6. Summary of findings in the present review**

**Supplementary Table S7. Mean or median birth weight of mothers with or without GDM in the studies that were not included in the meta-analysis**

**Supplementary Figure S1. Traffic light plot regarding risk of bias judgment in the included studies.**

The risk of bias were assessed in six domains (study participation, study attrition, prognostic factor measurement, outcome assessment, study confounding, and statistical analysis and reporting) and summarized as overall. The overall risk of bias was rated low if all QUIPS domains had a low risk of bias, moderate if there was one domain with an unclear or high risk of bias, and high if there were two or more domains with an unclear or high risk of bias.

**Supplementary Figure S2. Contour-enhanced funnel plot of estimates from the included studies**

The contour lines differentiate the significance and non-significance regions in the plot at 1%, 5% and 10% significance levels. The *x*-axis shows the effect-estimates (log-adjusted odds ratios) from the included studies; the *y*-axis shows the inverse of the log standard errors.

**Supplementary Figure S3. Sensitivity analysis of the association of pregnant women born low birth weight, small for gestational age, or preterm with gestational diabetes using crude odds ratios instead of adjusted odds ratios**

ES indicates effect size (crude odds ratio of each study); CI, confidence interval. The ES were determined using a random-effects meta-analysis model weighted by the inverse variance estimate. Square data markers represent ES, with marker size reflecting the statistical weight of the study using random-effects meta-analysis; horizontal lines, 95% CIs; diamond, the subgroup and overall odds ratios and 95% CIs for gestational diabetes

**Supplementary Figure S4.** **Sensitivity analysis of the association of pregnant women born low birth weight, small for gestational age, or preterm with gestational diabetes using crude odds ratios excluding studies without standard definition of gestational diabetes mellitus**

ES indicates effect size (adjusted odds ratio of each study); CI, confidence interval. The ES were determined using a random-effects meta-analysis model weighted by the inverse variance estimate. Square data markers represent ES, with marker size reflecting the statistical weight of the study using random-effects meta-analysis; horizontal lines, 95% CIs; diamond, the subgroup and overall odds ratios and 95% CIs for gestational diabetes.

**Supplementary Figure S5. Sensitivity analysis of the association of pregnant women born low birth weight, small for gestational age, or preterm with gestational diabetes excluding women born with macrosomia**

ES indicates effect size (adjusted odds ratio of each study); CI, confidence interval. The ES were determined using a random-effects meta-analysis model weighted by the inverse variance estimate. Square data markers represent ES, with marker size reflecting the statistical weight of the study using random-effects meta-analysis; horizontal lines, 95% CIs; diamond, the subgroup and overall odds ratios and 95% CIs for gestational diabetes.

**Supplementary Table S1**. MOOSE checklist for the present review

| **Item No** | **Recommendation** | **Reported on Page No** |
| --- | --- | --- |
| Reporting of background should include | | |
| 1 | Problem definition | 5 |
| 2 | Hypothesis statement | 5 |
| 3 | Description of study outcome(s) | 5 |
| 4 | Type of exposure or intervention used | 5 |
| 5 | Type of study designs used | 5 |
| 6 | Study population | 5 |
| Reporting of search strategy should include | | |
| 7 | Qualifications of searchers (eg, librarians and investigators) | 6 |
| 8 | Search strategy, including time period included in the synthesis and key words | 6, Table S2 |
| 9 | Effort to include all available studies, including contact with authors | 6-7 |
| 10 | Databases and registries searched | 6 |
| 11 | Search software used, name and version, including special features used (eg, explosion) | 6, Table S2 |
| 12 | Use of hand searching (eg, reference lists of obtained articles) | 6 |
| 13 | List of citations located and those excluded, including justification | Table S3 |
| 14 | Method of addressing articles published in languages other than English | 6 |
| 15 | Method of handling abstracts and unpublished studies | 6 |
| 16 | Description of any contact with authors | 7 |
| Reporting of methods should include | | |
| 17 | Description of relevance or appropriateness of studies assembled for assessing the hypothesis to be tested | 6-8 |
| 18 | Rationale for the selection and coding of data (eg, sound clinical principles or convenience) | 6-8 |
| 19 | Documentation of how data were classified and coded (eg, multiple raters, blinding and interrater reliability) | 6-8 |
| 20 | Assessment of confounding (eg, comparability of cases and controls in studies where appropriate) | 8 |
| 21 | Assessment of study quality, including blinding of quality assessors, stratification or regression on possible predictors of study results | 7-8 |
| 22 | Assessment of heterogeneity | 8 |
| 23 | Description of statistical methods (eg, complete description of fixed or random effects models, justification of whether the chosen models account for predictors of study results, dose-response models, or cumulative meta-analysis) in sufficient detail to be replicated | 8-9 |
| 24 | Provision of appropriate tables and graphics | Figure 1-3 |
| Reporting of results should include | | |
| 25 | Graphic summarizing individual study estimates and overall estimate | Figs 2-3 |
| 26 | Table giving descriptive information for each study included | Table S4 |
| 27 | Results of sensitivity testing (eg, subgroup analysis) | Figure S3-S5 |
| 28 | Indication of statistical uncertainty of findings | 10, Table S6 |

| **Item No** | **Recommendation** | **Reported on Page No** |
| --- | --- | --- |
| Reporting of discussion should include | | |
| 29 | Quantitative assessment of bias (eg, publication bias) | 11-13 |
| 30 | Justification for exclusion (eg, exclusion of non-English language citations) | 11 |
| 31 | Assessment of quality of included studies | 12 |
| Reporting of conclusions should include | | |
| 32 | Consideration of alternative explanations for observed results | 12 |
| 33 | Generalization of the conclusions (ie, appropriate for the data presented and within the domain of the literature review) | 13-14 |
| 34 | Guidelines for future research | 12 |
| 35 | Disclosure of funding source | 15 |

*From*: Stroup DF, Berlin JA, Morton SC, et al, for the Meta-analysis Of Observational Studies in Epidemiology (MOOSE) Group. Meta-analysis of Observational Studies in Epidemiology. A Proposal for Reporting. *JAMA*. 2000;283(15):2008-2012. doi: 10.1001/jama.283.15.2008.

**Supplementary Table S2**. Search strategies

| Medline via Ovid |  |
| --- | --- |
| 1 | exp infant, low birth weight/ or exp infant, small for gestational age/ or exp infant, premature/ |
| 2 | (low birth* or preterm birth*).tw. |
| 3 | exp pregnant women/ or exp pregnancy/ |
| 4 | pregnan*.tw. |
| 5 | (preeclampsia or eclampsia or gestational hypertension or HELLP or gestational diabetes or diabetes during pregnancy or GDM or pregnancy diabetes).tw. |
| 6 | exp Pre-Eclampsia/ or exp Eclampsia / or exp HELLP syndrome/ or Diabetes, Gestational/ |
| 7 | 1 or 2 |
| 8 | 3 or 4 |
| 9 | 5 or 6 |
| 10 | 7 and 8 and 9 |
| EMBASE via ProQuest |  |
| 1 | ab(low birth*) OR ti(low birth*) |
| 2 | ab(preterm birth*) OR ti(preterm birth*) |
| 3 | EMB.EXACT.EXPLODE ("low birth weight") OR EMB.EXACT.EXPLODE ("small for date infant") OR EMB.EXACT.EXPLODE("prematurity") |
| 4 | 1 OR 2 OR 3 |
| 5 | ab(pregnan*) OR ti(pregnan*) |
| 6 | EMB.EXACT.EXPLODE("pregnant woman") OR EMB.EXACT.EXPLODE("pregnancy") |
| 7 | 5 OR 6 |
| 8 | ab(preeclampsia) OR ti(preeclampsia) |
| 9 | ab(gestational hypertension) OR ti(gestational hypertension) |
| 10 | ab(HELLP) OR ti(HELLP) |
| 11 | ab(gestational diabetes) OR ti(gestational diabetes) |
| 12 | ab(diabetes during pregnancy) OR ti(diabetes during pregnancy) |
| 13 | ab(pregnancy diabetes) OR ti(pregnancy diabetes) |
| 14 | ab(GDM) OR ti(GDM) |
| 15 | EMB.EXACT.EXPLODE ("eclampsia and preeclampsia") OR EMB.EXACT.EXPLODE("HELLP syndrome") OR EMB.EXACT.EXPLODE("pregnancy diabetes mellitus") |
| 16 | 8 OR 9 OR 10 OR 11 OR 12 OR 13 OR 14 OR 15 |
| 17 | 4 AND 7 AND 16 |
| EBSCO CINAHL |  |
| 1 | MH infant, low birth weight OR MH infant, small for gestational age OR MH infant, premature |
| 2 | AB low birth* OR AB preterm birth* |
| 3 | MH pregnant women OR MH pregnancy |
| 4 | AB pregnan* |
| 5 | AB preeclampsia OR AB eclampsia OR AB gestational hypertension OR AB HELLP OR AB gestational diabetes OR AB diabetes during pregnancy OR AB GDM OR AB pregnancy diabetes |
| 6 | MH Pre-Eclampsia OR MH Eclampsia OR MH HELLP syndrome OR MH Diabetes, Gestational |
| 7 | S1 OR S2 |
| 8 | S3 OR S4 |
| 9 | S5 OR S6 |
| 10 | S7 AND S8 AND S9 |
| ClinicalTrials.gov |  |
| Study type: | Observational Studies \| |
| Condition or disease: | weight OR preterm |
| Other terms: | preeclampsia OR eclampsia OR gestational hypertension OR complications |
| The world health organization international clinical trials platform search portal (WHO-ICTRP) | |
| 1 | Title (weight OR preterm) AND (preeclampsia OR eclampsia OR gestational hypertension OR complications) |
| 2 | Condition (weight OR preterm) AND (preeclampsia OR eclampsia OR gestational hypertension OR complications) |
| 3 | #1 OR #2 |

**Supplementary Table S3**. List of studies that were excluded from this review and the reason for exclusion

| References | Reason for exclusion |
| --- | --- |
| Bonamy A-KE, Norman M, Kaijser M. Being born too small, too early, or both: Does it matter for risk of hypertension in the elderly? American Journal of Hypertension. 2008;21(10):1107-10. | Wrong participants |
| Carr H, Cnattingius S, Granath F, Ludvigsson JF, Edstedt Bonamy AK. Preterm Birth and Risk of Heart Failure Up to Early Adulthood. J Am Coll Cardiol. 2017;69(21):2634-42. | Wrong participants |
| Carr H, Cnattingius S, Ludvigsson JF, Bonamy A-KE. Risk of heart failure in children and young adults born preterm-a national cohort study. Circulation. 2015;132. | Wrong participants |
| Crump C, Howell EA, Stroustrup A, McLaughlin MA, Sundquist J, Sundquist K. Association of Preterm Birth With Risk of Ischemic Heart Disease in Adulthood. JAMA Pediatr. 2019. | Wrong participants |
| Dempsey JC, Williams MA, Luthy DA, Emanuel I, Shy K. Weight at birth and subsequent risk of preeclampsia as an adult. American journal of obstetrics and gynecology. 2003;189(2):494-500. | Wrong Outcomes |
| Hennessy E, Alberman E. Intergenerational influences affecting birth outcome. II. Preterm delivery and gestational age in the children of the 1958 British birth cohort. Paediatric and Perinatal Epidemiology. 1998;12:61-75. | Wrong participants |
| Hofman PL, Cutfield WS. Insulin sensitivity in people born pre-term, with low or very low birth weight and small for gestational age. Journal of endocrinological investigation. 2006;29(1):2-8. | Wrong publication type |
| Hovi P, Vohr B, Ment LR, Doyle LW, McGarvey L, Morrison KM, et al. Blood Pressure in Young Adults Born at Very Low Birth Weight: Adults Born Preterm International Collaboration. Hypertension. 2016;68(4):880-7. | Wrong participants |
| Keijzer-Veen MG, Dülger A, Dekker FW, Nauta J, Van Der Heijden BJ. Very preterm birth is a risk factor for increased systolic blood pressure at a young adult age. Pediatric Nephrology. 2010;25(3):509-16. | Wrong participants |
| Klebanoff MA, Secher NJ, Mednick BR, Schulsinger C. Maternal size at birth and the development of hypertension during pregnancy: A test of the Barker hypothesis. Archives of Internal Medicine. 1999;159(14):1607-1612 | Wrong outcomes |
| Kurabayashi T, Mizunuma H, Kubota T, Nagai K, Hayashi K. Low Birth Weight and Prematurity Are Associated with Hypertensive Disorder of Pregnancy in Later Life: A Cross-Sectional Study in Japan. American journal of perinatology. 2020. | Wrong outcomes |
| Lawlor DA, Ronalds G, Clark H, Smith GD, Leon DA. Birth weight is inversely associated with incident coronary heart disease and stroke among individuals born in the 1950s: Findings from the Aberdeen children of the 1950s prospective cohort study. Circulation. 2005;112(10):1414-8. | Wrong participants |
| Lewandowski AJ, Augustine D, Lamata P, Davis EF, Lazdam M, Francis J, et al. Preterm heart in adult life: Cardiovascular magnetic resonance reveals distinct differences in left ventricular mass, geometry, and function. Circulation. 2013;127(2):197-206. | Wrong participants |
| Lewandowski AJ, Augustine DX, Davis EF, Lazdam M, Banerjee R, Singhal A, et al. Unique left ventricular geometry and function of young adults born preterm: Impact of prematurity and preeclampsia exposure. European Heart Journal. 2012;33:1041. | Wrong participants |
| Lewandowski AJ, Davis EF, Singhal A, Lucas A, McCormick K, Shore AC, et al. Increased blood pressure in preterm-born individuals correlates with a distinct antiangiogenic state and microvascular abnormalities in adult life. Circulation. 2014;130. | Wrong participants |
| Lindström L, Skjærven R, Bergman E, Lundgren M, Klungsøyr K, Cnattingius S, et al. Chronic Hypertension in Women after Perinatal Exposure to Preeclampsia, Being Born Small for Gestational Age or Preterm. Paediatric and Perinatal Epidemiology. 2017;31(2):89-98. | Wrong outcomes |
| Luo ZC, Fraser WD, Julien P, Deal CL, Audibert F, Smith GN, et al. Tracing the origins of "fetal origins" of adult diseases: Programming by oxidative stress? Medical Hypotheses. 2006;66(1):38-44. | Wrong publication type |
| Machado Pereira SS, Jesus Oliveira MdN, Rodrigues Correia Koller JM, Amorim Miranda FC, Pires Ribeiro I, da Silva Oliveira AD. Profile of the Pregnant Women Affected by Preterm Birthin a Public Maternity Hospital. Revista de Pesquisa: Cuidado e Fundamental. 2018;10(3):758-63. | Wrong participants |
| Miettola S, Hovi P, Andersson S, Strang-Karlsson S, Pouta A, Laivuori H, et al. Maternal preeclampsia and bone mineral density of the adult offspring. American Journal of Obstetrics and Gynecology. 2013;209(5):443.e1-.e10. | Wrong participants |
| Nahum Sacks K, Friger M, Shoham-Vardi I, Spiegel E, Sergienko R, Landau D, et al. Prenatal exposure to preeclampsia as an independent risk factor for long-term cardiovascular morbidity of the offspring. Pregnancy Hypertension. 2018;13:181-6. | Wrong participants |
| Näsänen-Gilmore SPK, Sipola-Leppänen M, Tikanmäki M, Miettola S, Matinolli HM, Turkka S, et al. Effect of preterm birth on adulthood lung function: Finnish birth registry and clinical follow-up study. Archives of Disease in Childhood. 2014;99:A67. | Wrong participants |
| Oxford Uo, Foundation BH. The Effect of Prematurity and Hypertensive Disorders of Pregnancy on Offspring Cardiovascular Health: https://ClinicalTrials.gov/show/NCT01888770; 2011 2011. | Wrong participants |
| Paz Levy D, Sheiner E, Wainstock T, Sergienko R, Landau D, Walfisch A. Evidence that children born at early term (37-38 6/7 weeks) are at increased risk for diabetes and obesity-related disorders. American Journal of Obstetrics and Gynecology. 2017;217(5):588.e1-.e11. | Wrong participants |
| Paz Y Miño F, Casu G, Crovetto F, Gratacós E, Crispi F, Sepúlveda‐Martínez Á, et al. Transgenerational transmission of small-for-gestational age. Ultrasound in Obstetrics & Gynecology. 2019;53(5):623-9. | Wrong participants |
| Persson JL, Stuart A, Amer-Wahlin I, Kallen K. Long-term cardiovascular risk in relation to birth weight and exposure to maternal diabetes mellitus. European Heart Journal. 2013;34:958-9. | Wrong participants |
| Pocobelli G, Dublin S, Enquobahrie DA, Mueller BA. Birth Weight and Birth Weight for Gestational Age in Relation to Risk of Hospitalization with Primary Hypertension in Children and Young Adults. Maternal and child health journal. 2016;20(7):1415-23. | Wrong participants |
| Qanitha A, De Mol BAJM, Pabittei DR, Mappangara I, Kabo P, Uiterwaal CSPM. Maternal pregnancy complications and premature coronary heart disease in the offspring. European Heart Journal. 2016;37:933. | Wrong participants |
| Rasmussen S, Irgens LM. Pregnancy-induced hypertension in women who were born small. Hypertension. 2007;49(4):806-812. | Wrong outcomes |
| Scholten RR, Krabbendam I, Oyen WJ, Hopman MT, Lotgering FK, Spaanderman ME. Women born small for gestational age have lower plasma volume in adult life. Reproductive Sciences. 2010;17(3):357A-8A. | Wrong participants |
| Sepúlveda-Martínez Á, Rodríguez-López M, Paz YMF, Casu G, Crovetto F, Gratacós E, et al. Transgenerational transmission of small-for-gestational age. Ultrasound Obstet Gynecol. 2019;53(5):623-9. | Wrong participants |
| Sherf Y, Sheiner E, Shoham Vardi I, Sergienko R, Klein J, Bilenko N. 452: Recurrence of preterm delivery in women with a family history of preterm delivery. American Journal of Obstetrics and Gynecology. 2016;214(1). | Wrong outcomes |
| Sherf Y, Sheiner E, Vardi IS, Bilenko N. Like mother like daughter-low birth weight and preeclampsia tend to re-ocurre at the next generation. American Journal of Obstetrics and Gynecology. 2015;212(1):S34. | Wrong outcomes |
| Sherf Y, Sheiner E, Vardi IS, Sergienko R, Klein J, Bilenko N. Recurrence of Preterm Delivery in Women with a Family History of Preterm Delivery. American Journal of Perinatology. 2017;34(4):397-402. | Wrong outcomes |
| Sherf Y, Shoham Vardi I, Sergienko R, Bilenko N, Sheiner E, Klein J. Like mother like daughter: low birth weight and preeclampsia tend to reoccur at the next generation. Journal of Maternal-Fetal & Neonatal Medicine. 2019;32(9):1478-84. | Wrong outcomes |
| Wagata M, Tsuchiya N, Nakaya N, et al. The association between woman's own birth weight and her subsequent risk for hypertensive disorders of pregnancy. Reproductive Sciences. 2018;25(1):240A | Wrong outcomes |
| Wikstrom AK, Svensson T, Kieler H, Cnattingius S. Recurrence of placental dysfunction disorders across generations. Am J Obstet Gynecol. 2011;205(5):454 e451-458 | Wrong outcomes |
| Zetterstrom K, Lindeberg S, Haglund B, Magnuson A, Hanson U. Being born small for gestational age increases the risk of severe pre-eclampsia. BJOG : an international journal of obstetrics and gynaecology. 2007;114(3):319-324. | Wrong outcomes |

Supplementary Table S4. Detail characteristics of the included studies

| **Study** | **Methods** | **Subject characteristics** | **Exposure and control used in the present review** | **Gestational diabetes mellitus definition** | **Adjusted confounders in the present review** | **Notes** |
| --- | --- | --- | --- | --- | --- | --- |
| a ́ Rogvi 2012 | Design: retrospective cohort Setting: population-based Location: Denmark Sample size: 116,595 | Age: 24.7 ± 2.8 Birth year: 1978 to 1981 Ethnicity: not reported Smoking: not reported Primiparous: not reported | Exposure: gestational age <37 weeks (n = 1329)  Control: gestational age ≥37 weeks (n = 31,047) | GDM defined by ICD-8: 634.74 or ICD-10: O24.4 | Unadjusted | Published data only |
| Andraweera 2019 | Design: prospective cohort Setting: hospital-based (multicentre) Location: international Sample size: 5,327 | Age: 28.7 ± 5.4 Birth year: not reported Ethnicity: non-Hispanic White, Asian, Polynesian, Indian, others  Smoking: 566 (10.1)  Primiparous: 5327 (100) | Exposure: birth weight of <2500g (n = not reported)  Control: birth weight of ≥2500g (n = not reported) | GDM defined by fasting glucose ≥ 5.1 mmol/L or a 2-hour level of ≥ 8.5 mmol/L following an oral glucose tolerance test, according to the new WHO classification | Maternal age, smoking at 15 weeks’ gestation, ethnicity, socioeconomic status, family history of diabetes, maternal gestational age at birth, infant sex, and recruitment centre. | Unpublished and published data |
| Bo 2003 | Design: case control Setting: hospital-based (single centre) Location: Italy Sample size: 300 | Age: not reported Birth year: not reported Ethnicity: not reported Smoking: 100 (33.3) Primiparous: not reported | Birth weight as a continuous variable | GDM defined by fasting glucose ≥ 95 mg/dl, a 2-hour level of ≥ 155 mg/dl, or a 3-hour level of ≥ 140 mg/dl.  Impaired glucose tolerance defined that only one glucose value was higher than above cut-off levels. | Maternal diabetes | Published data only |
| Boivin 2012 | Design: retrospective cohort Setting: population-based Location: Canada Sample size: 24,119 | Age: Preterm, 23.1 ± 3.7; Term 23.4 ± 3.8 Birth year: 1976 to 1995 Ethnicity: not reported Smoking: not reported Primiparous: 12,130 (50.3) | Exposure: gestational age of 32 to 36 weeks (n = 6851)  Control: gestational age of 37 to 42 weeks (n = 16,714) | GDM defined by ICD-9 before April 1, 2006, and the ICD-10 after April 1, 2006. | Mother’s birth characteristics (SGA, large for gestational age, multiple births, and year of birth), chronic hypertension, diabetes, kidney disease, age ≥ 25 years and multiple-birth pregnancy. | Unpublished and published data |
| Chawla 2014 | Design: retrospective cohort Setting: population-based Location: US Sample size: 130,617 | Age: mainly 25 to 35 Birth year: 1956 to 1976 Ethnicity: non-Hispanic White, Hispanic, and African Smoking: not reported Primiparous: 60,958 (46.7) | Exposure: SGA (n = 13,934)  Control: AGA (n = 116,658) | Diabetes during pregnancy defined as pre-existing DM (including  monogenic, Type 1 and Type 2) and gestational DM. | Maternal age, education, parity, plurality, marital status, and race/ethnicity | Published data only |
| Crusell 2017 | Design: case control Setting: hospital-based (single centre) Location: Denmark Sample size: 1,322 | Age: mainly 26 to 28 Birth year: 1939 to 1970 Ethnicity: not reported Smoking: not reported Primiparous: not reported | Birth weight as a continuous variable | GDM based on local criteria and changed during the period: until 1987 a 3 h 50 g OGTT, and after 1987 a 3 h 75 g OGTT. GDM was diagnosed when two or more out of seven measurements exceeded the mean +3 standard deviations on a curve based on non-pregnant normal-weight women without a family history of diabetes | Unadjusted | Published data only |
| Egeland 2000 | Design: retrospective cohort Setting: population-based Location: Norway Sample size: 138,714 | Age: 14 to 31 Birth year: 1967 to 1984 Ethnicity: not reported Smoking: not reported Primiparous: not reported | Exposure: birth weight of <2500g (n = 4,652)  Control: birth weight of 4000 to 4500g (n = 14,852) | Self-reported GDM in one or more pregnancies | Women’s age and parity and their mother’s diabetic status. | Published data only |
| Innes 2003 | Design: case control Setting: population-based Location: USA Sample size: 23,395 | Age: mainly 17 to 24 Birth year: 1970 or later Ethnicity: White, non-Hispanic, Hispanic, other non-White Smoking: 4,258 (18.2) Primiparous: 23,395 (100) | Exposure: birth weight of 2000 to 2499 g (n = 1325)  Control: birthweight of 3500g to 3999 g (n = 5639) | GDM defined by ICD-9 code 648.0 or abnormal glucose tolerance defined by ICD-9 code 648.8 on their hospital discharge records. | Age, race, education, employment status, pre-pregnancy body mass index, height, pregnancy weight gain, and gestational age, | Published data only |
| Legarros 2012 | Design: retrospective cohort  Setting: population-based Location: Sweden Sample size: 323,083 | Age: mainly 20 to 29 Birth year: 1973 or later Ethnicity: not reported Smoking: 43,397 (13.4) Primiparous: 196,859 (60.9) | Exposure: SGA defined as more than 2 SD below the mean birth-weight-for-gestational-age (n = 12,083)  Control: Birth weight 1 SD below the mean to 1 SD above the mean (n = 214,905) | GDM defined by the ICD-9 code 648W and the ICD-10 code O244, those diagnosis were mainly based on a 75 g oral glucose tolerance test with a fasting capillary whole blood glucose level C 6.1 mmol/L (plasma C 7.0 mmol/L) and/or a 2 h blood glucose C 9.0 mmol/L (plasma glucose C 10.0 mmol/L) | Body mass index height, maternal age, education, parity, smoking, and year of pregnancy | Published data only |
| Moses 1999 | Design: case control Setting: hospital-based (single centre) Location: Australia Sample size: 138 | Age: case, 26 (5.8); control, 26 (5.6) Birth year: not reported Ethnicity: not reported Smoking: not reported Primiparous: not reported | Birth weight as a continuous variable | GDM defined by fasting glucose is ≥5.5 mmol/l and/or the 2-h glucose level is ≥8.0 mmol/l, according to the criteria of the Australasian Diabetes in Pregnancy Society. | Unadjusted | Published data only |
| Ogonowski 2014 | Design: case control Setting: hospital-based (multicentre) Location: Poland Sample size: 1,588 | Age: 29.7 (0.6) Birth year: not reported Ethnicity: Caucasian Smoking: not reported Primiparous: 902 (56.8) | Exposure: birth weight of < 2500g (n = not reported)  Control: birth weight of 3500 to 3999g (n = not reported) | GDM defined by either the fasting glucose was 7.0 mmol/L or the 2-h glucose concentration was 7.8 mmol/L, which was in accordance with the WHO diagnostic criteria | Age, body mass index, family history of diabetes, and prior gestational diabetes. | Published data only |
| Olah 1996 | Design: retrospective cohort Setting: hospital-based (single centre) Location: UK Sample size: 592 | Age: 27 (range 19 to 42) Birth year: not reported Ethnicity: predominantly Caucasian Smoking: not reported Primiparous: not reported | Exposure: birth weight of <2500g (n = 57)  Control: birth weight of ≥ 2500g (n = 452) | Impaired glucose tolerance, defined as a 2-hour plasma glucose concentration of 7.8-1 1.0 mmol/L, or gestational diabetes, defined as a 2-hour plasma glucose concentration of 11.1 mmol/L or more. | Unadjusted | Published data only |
| Pettit 1998 | Design: retrospective cohort Setting: not reported Location: US Sample size: 831 | Age: not reported Birth year: not reported Ethnicity: Pima Indians Smoking: not reported Primiparous: not reported | Exposure: birth weight of <2500g (n = 29)  Control: birth weight of ≥2500g (n = 802) | Diabetes during pregnancy diagnosed by the WHO criteria, i.e., a glucoses concentration of ≥ 11.1 mmol/L 2 h after the ingestion of a 75-g glucose load | Unadjusted | Published data only |
| Plante 1999 | Design: retrospective cohort Setting: population-based Location: US Sample size: 6,767 | Age: 19 to 22 Birth year: 1974 Ethnicity: non-Hispanic White, African Smoking: not reported Primiparous: not reported | Exposure: SGA defined as a birth weight less than the tenth percentile for gestational age, race-specific for either black or white (n = 596)  Control: not SGA (n = 5,954) | Diabetes on the birth certificate records. The coding was performed by the physician or other individual filling out the birth record and consisted of a checkbox in the field described as “medical complications of pregnancy.” | Unadjusted | Published data only |
| Plante 2004 | Design: retrospective cohort Setting: population-based Location: US Sample size: 7,802 | Age: 24 to 26 Birth year: 1974 Ethnicity: non-Hispanic White, African Smoking: not reported Primiparous: not reported | Exposure: SGA as defined in the Plante 1999 study (n = 537)  Control: AGA as defined in the Plante 1999 study (n = 7,265) | Diabetes on the birth certificate. Specific information as to criteria for diagnosis was not available | Unadjusted | Published data only |
| Savona-Ventura 2003 | Design: case control Setting: hospital-based (single centre) Location: Malta Sample size: 7,075 | Age: not reported Birth year: 1952 to 1983 Ethnicity: not reported Smoking: not reported Primiparous: not reported | Exposure: birth weight of <2500g (n = 419)  Control: birth weight of ≥2500g (n = 6,656) | GDM defined by a serum glucose concentration >8.6 mmol/l 2 hours after the OGTT. | Unadjusted | Published data only |
| Seghieri 2002 | Design: retrospective cohort Setting: hospital-based (single centre) Location: Italy Sample size: 604 | Age: LBW: 31.7 (4.2)) Birth year: not reported Ethnicity: not reported Smoking: not reported Primiparous: not reported | Exposure: birth weight of <2500g (n = 68)  Control: birth weight of ≥2500g (n = 536) | GDM defined by the American Diabetes Association | Age, parity, family history of diabetes, and pre-pregnancy BMI, | Published data only |
| Williams 1999 | Design: retrospective cohort Setting: population-based Location: US Sample size:41,839 births (38,513 mothers) | Age: mainly 19 to 29 Birth year: 1949 to 1979 Ethnicity: non-Hispanic White, Hispanic, African, native American Smoking: 10,429 (24.9) Primiparous: 32,488 (77.7) | Exposure: birth weight of <2500g (n = 2,708 births)  Control: birth weight of ≤2500g (n = 39,131 births) | GDM recorded on the birth certificate and/or given an ICD-9 diagnosis | Unadjusted | Published data only |
| Data are presented as the mean ± SD or number (percentage).  GDM, gestational diabetes mellitus, ICD, International Classification of Diseases; WHO, World Health Organization; SGA, small for gestational age; AGA, appropriate for gestational age; SD, standard deviation; OGTT, oral glucose tolerance test | | | | | | |

**Supplementary Table S5**. Details of inclusion and exclusion criteria for the original studies

| Study ID | Inclusion criteria | Exclusion criteria |
| --- | --- | --- |
| a Rogvi 2012 ^1^ | Women with a registered childbirth in the years 1989–2007, and born between 1974 to 1981 in the Danish Medical Birth Registry | Not reported |
| Andraweera 2019 ^2^ | Nulliparous women with singleton pregnancies were recruited before 15 weeks’ gestation. | Those considered at high risk of preeclampsia, SGA, or preterm birth because of underlying medical conditions, gynecological history, or three or more miscarriages or terminations of pregnancy or couples who received medical or surgical interventions that could modify pregnancy outcome were not eligible |
| Bo 2003 ^3^ | Pregnant women born in Turin were included. The first consecutive 200 women with singleton pregnancy and normal glucose tolerance. 50 with ICT and 50 with GMD, identified by sequential screening, were selected. | Women known to have diabetes mellitus or a disease affecting glucose metabolism were excluded. |
| Boivin 2012 ^4,5^ | Exposed cohort: women born preterm between 1976 and 1995 in Quebec. Unexposed cohort: Women born at term at 37–42 weeks’ gestation | Women born before 23 weeks’ gestation and those born to multiple-birth (≥ 3) pregnancies. Women born at ≥ 43 weeks |
| Chawla 2014 ^6,7^ | Women between the ages of 15 and 35, who were born in Illinois between 1956 and 1976 and were non-Latina White, African-American, and Mexican-American. Only women with the first singleton gestations during the period between 1989 and 1991 | Women born large for gestational age and with missing gestational age |
| Crusell 2017 ^8^ | Women with previous diet treated GDM at the Center for Pregnant Women with Diabetes, Department of Obstetrics, Rigshospitalet, Copenhagen, Denmark, and who had willingness to follow-up. (Cases)  A control cohort was selected from a population-based non-pharmacological prevention study (Inter99) on ischaemic heart disease. This study was conducted around the same time as the follow-up study of the GDM cohort at the Research Centre for Prevention and Health, Glostrup, Denmark. The women in the control cohort were invited stratified on age: 1939–1940, 1944–1945, 1949–1950, 1954–1955, 1959–1960, 1964–1965 or 1969–1970. (Controls) | Women from the original cohort were excluded from this study if they did not meet the inclusion criteria: Danish parental ethnicity, born in Denmark as a singleton, and alive and still living in Denmark at the time of conducting this study |
| Egeland 2000 ^9^ | All women born in 1967-84 who had given birth between 1988 and 1998. | Women who were not singletons. |
| Innes 2003 ^10,11^ | All women who completed a first pregnancy (and delivered a liveborn infant) in Upstate New York between 1994 and 1998, and who were also born in New York State in 1970 or later. | Any women whose first pregnancies were complicated by multifetal gestation, by the use of illegal drugs, or by preexisting diabetes mellitus, heart disease, essential hypertension, renal disease, or other preexisting chronic or serious acute serious acute conditions requiring medication or monitoring. Women with pregnancy induced hypertension were also excluded. |
| Legarros 2012 ^12^ | Women who were included in the Birth Register both as infants and mothers between 1973 and 2006. For the second generation, we only included singleton births in 1992 or later. Further, only mothers who were born in singleton pregnancies themselves, of a mother without diabetes type I at the time of pregnancy (identified by the ICD-8 code 648A and ICD-9 code 250). | Pregnant mothers with pregestational diabetes (identified at first antenatal visit and/or by ICD-9 code 648A and ICD-10 code O2403) |
| Moses 1999 ^13^ | Cases: consecutive women who had a singleton pregnancy delivery at 37–41 weeks, and referred for the medical management of their GDM between January 1990 and June 1997.  Controls: the same gestational age from a mother with an age within 2 years of the mother with GDM. | Not reported |
| Ogonowski 2014 ^14^ | Cases: women with abnormal glucose challenge test results who were referred to the Outpatient Clinic for Diabetic Pregnant Women in Szczecin from 2000 to 2007 and were diagnosed as GDM.  Controls: healthy pregnant women who were  randomly selected from among women who had a previous pregnancy in the Department for Feto-Maternal Medicine in Szczecin. All of these women had normal GCT results during their initial visits and had no GDM until delivery. | Not reported |
| Olah 1996 ^15^ | Women who had glucose tolerance test at Liverpool Maternity Hospital during the period January 1992 to December 1993. | Women born before 37 weeks of gestation, and women who did not know and could not find out their birth weight. |
| Pettit 1998 ^16,17^ | Pima Indians at least 5 years of age and living in the Gila River Indian Community of Central Arizona | Not reported |
| Plante 1999 ^18^ | Women who were born in 1974 at 37–44 weeks’ gestation in the state of Pennsylvania and delivered babies in 1995 or 1996. Females born small for gestational age or appropriate for gestational age. | Multiple births and incomplete information were excluded. Mother who was a twin or was of racial categories other than black or white were excluded. |
| Plante 2004 ^19^ | All females born at term in Pennsylvania in 1974 who delivered singleton full-term live-born infants in the state between January 1 1999 and 31 December 2000. Females born small for gestational age or appropriate for gestational age. | Multiple births and incomplete information were excluded. Mother who was a twin or was of racial categories other than black or white were excluded. |
| Savona-Ventura 2003 ^20^ | Cases: women diagnosed as suffering from gestational diabetes mellitus in 1996–2000 and delivered in the government-run hospital  Controls: the total population born during the same period, calculated from previously published birth weight distribution pattern studies carried out in 1965 and 1981 | Not reported |
| Seghieri 2002 ^21^ | All women who are classified as glucose intolerant to a previous 1-h 50-g OGTT, who show a 1-h plasma glucose level 7.8 mmol/l, or who have other risk factors for GDM (history of glucose intolerance, of macrosomic deliveries during previous pregnancies, or of diabetes in first-degree relatives). | Not reported |
| Williams 1999 ^22,23^ | Pregnant women, born in Washington State since 1949, with singleton births occurring between 1987 and 1995 | Mothers who were twins or of other multiple birth, mothers noted to have established diabetes before the index pregnancy, and　women whose diagnosis of diabetes could not be determined to be pregnancy associated. |

| **Supplementary Table S6**. Summary of findings in the present review | | | | | | |
| --- | --- | --- | --- | --- | --- | --- |
| Outcome | Anticipated absolute effects^*^ (95% CI) | | Relative effect (95% CI) | № of participants  (studies) | Certainty of the evidence (GRADE) | Comments |
|  | Risk with those not | Risk with women born small or premature |  |  |  |  |
| Gestational diabetes | Low prevalence setting | | OR 1.84 (1.54 to 2.20) | 825,622 (15 observational studies) | ⨁⨁◯◯ LOW ^a,b^ | a. Due to high risk of bias  b. Due to funnel plot asymmetry |
|  | 20 per 1,000 | 36 per 1,000 (30 to 43) |  |  |  |  |
|  | Moderate prevalence setting | |  |  |  |  |
|  | 100 per 1,000 | 170 per 1,000 (146 to 196) |  |  |  |  |
|  | High prevalence setting | |  |  |  |  |
|  | 200 per 1,000 | 315 per 1,000 (278 to 355) |  |  |  |  |
| *The risk in the intervention group (and its 95% confidence interval) is based on the assumed risk in the comparison group and the relative effect of the intervention (and its 95% CI).   CI: Confidence interval; OR: Odds ratio | | | | | | |
| GRADE Working Group grades of evidence High certainty: We are very confident that the true effect lies close to that of the estimate of the effect Moderate certainty: We are moderately confident in the effect estimate: The true effect is likely to be close to the estimate of the effect, but there is a possibility that it is substantially different Low certainty: Our confidence in the effect estimate is limited: The true effect may be substantially different from the estimate of the effect Very low certainty: We have very little confidence in the effect estimate: The true effect is likely to be substantially different from the estimate of effect | | | | | | |

| Table F. Mean or median birth weight of mothers with or without GDM | | |
| --- | --- | --- |
| Characteristics | Mothers with GDM | Mothers without GDM |
| Bo 2003 | 3077 ± 661 | 3389 ± 644 |
| Crusell 2017 | 3242 (3166 to 3318) | 3354 (3324 to 3384) |
| Moses 1999 | 3293 ± 493 | 3315 ± 460 |
| Numbers are displayed as mean ± SD or median (interquartile range). GDM, gestational diabetes mellitus. | | |

**Supplementary Figure S1**.

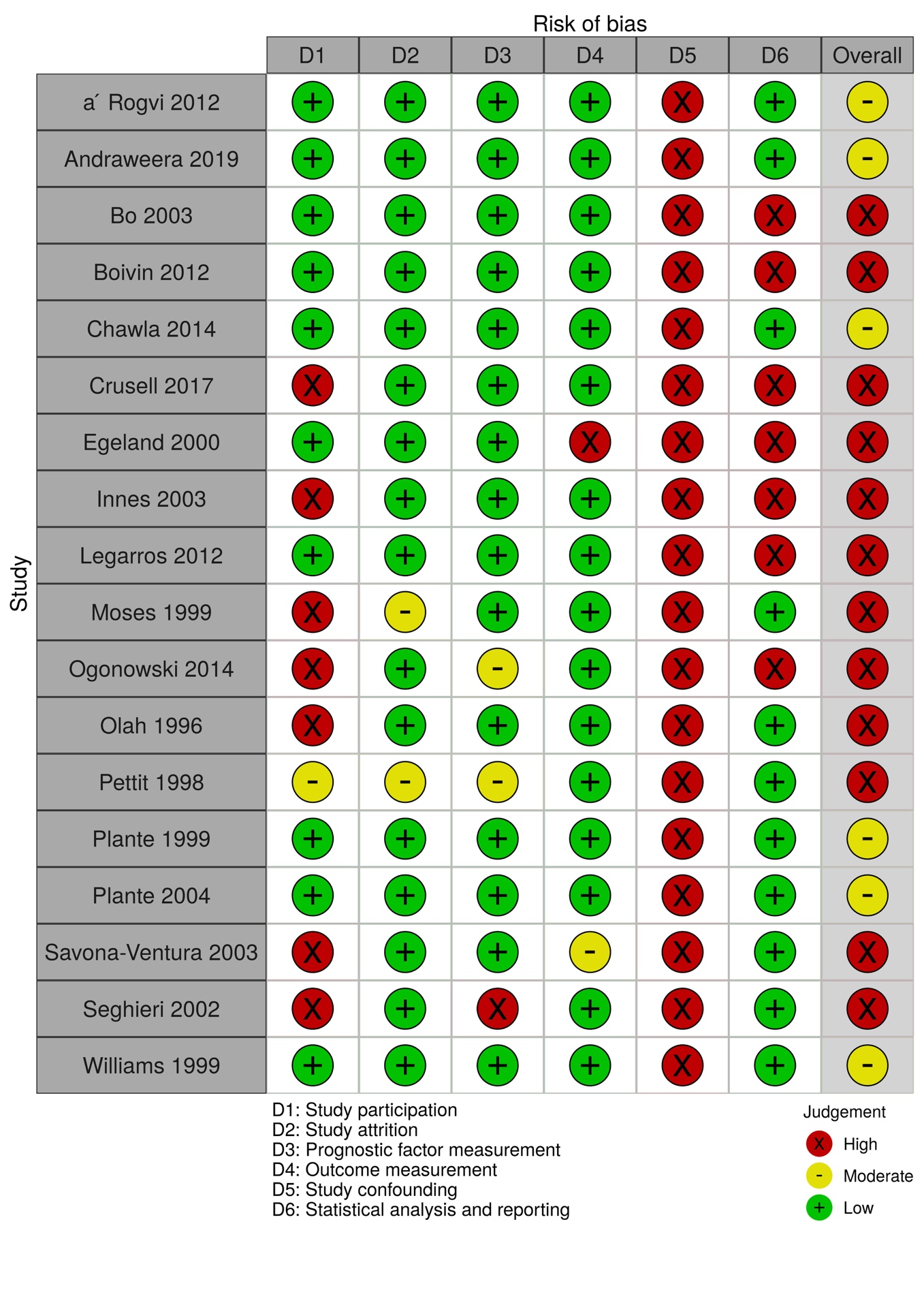

**Supplementary Figure S**2.

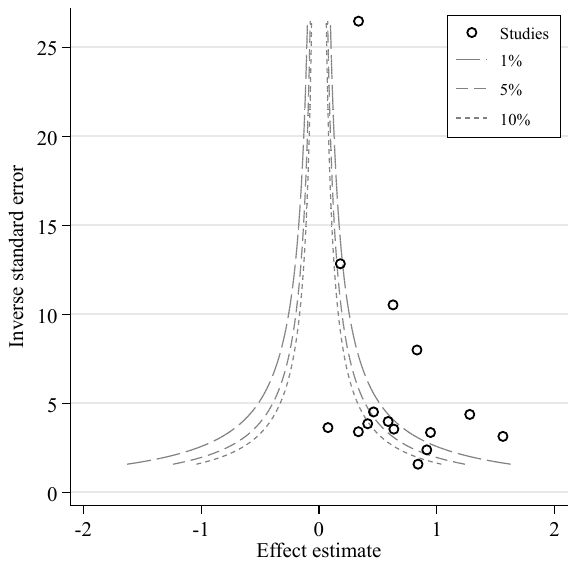

**Supplementary Figure S3**
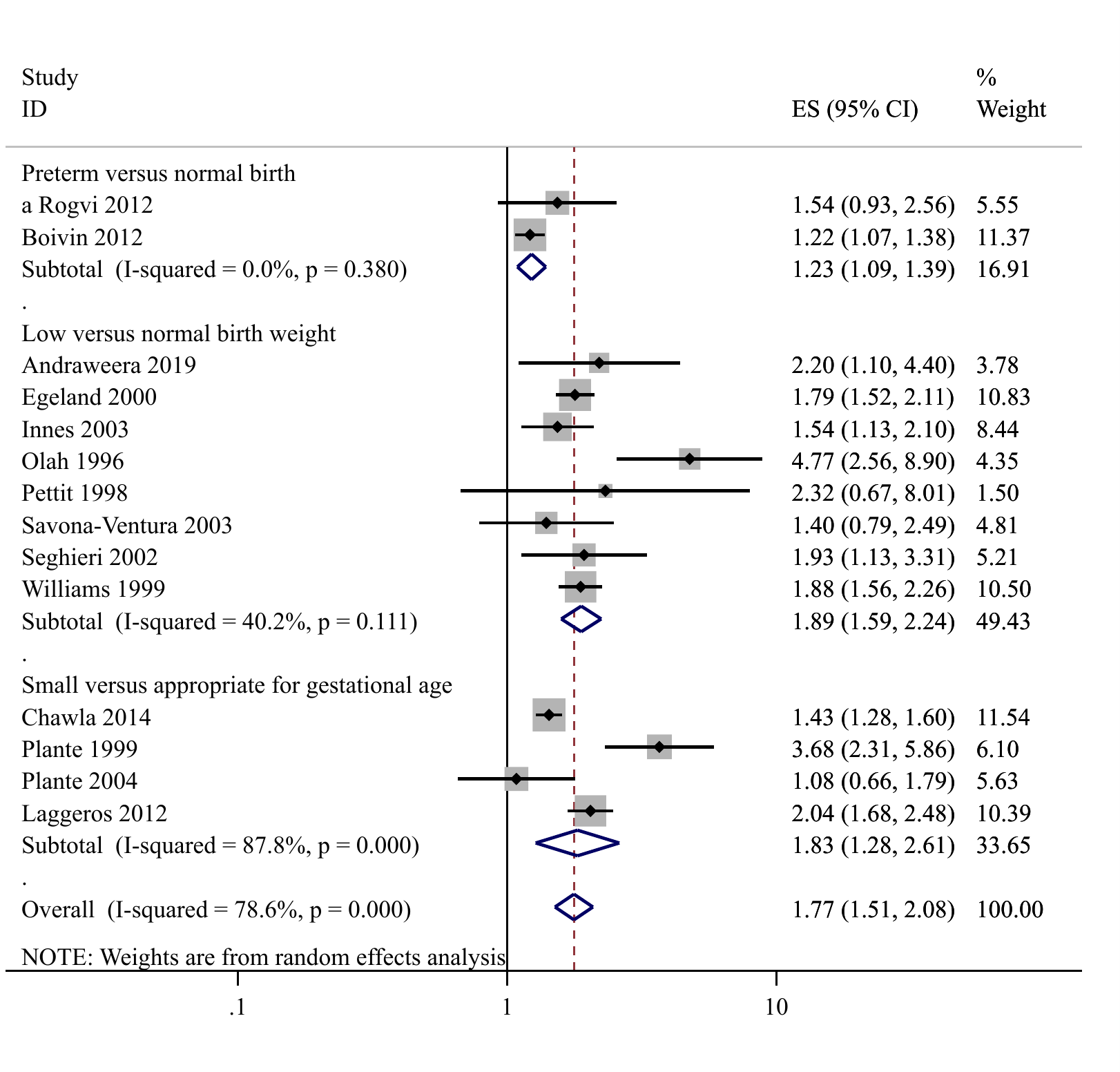

**Supplementary Figure S4**
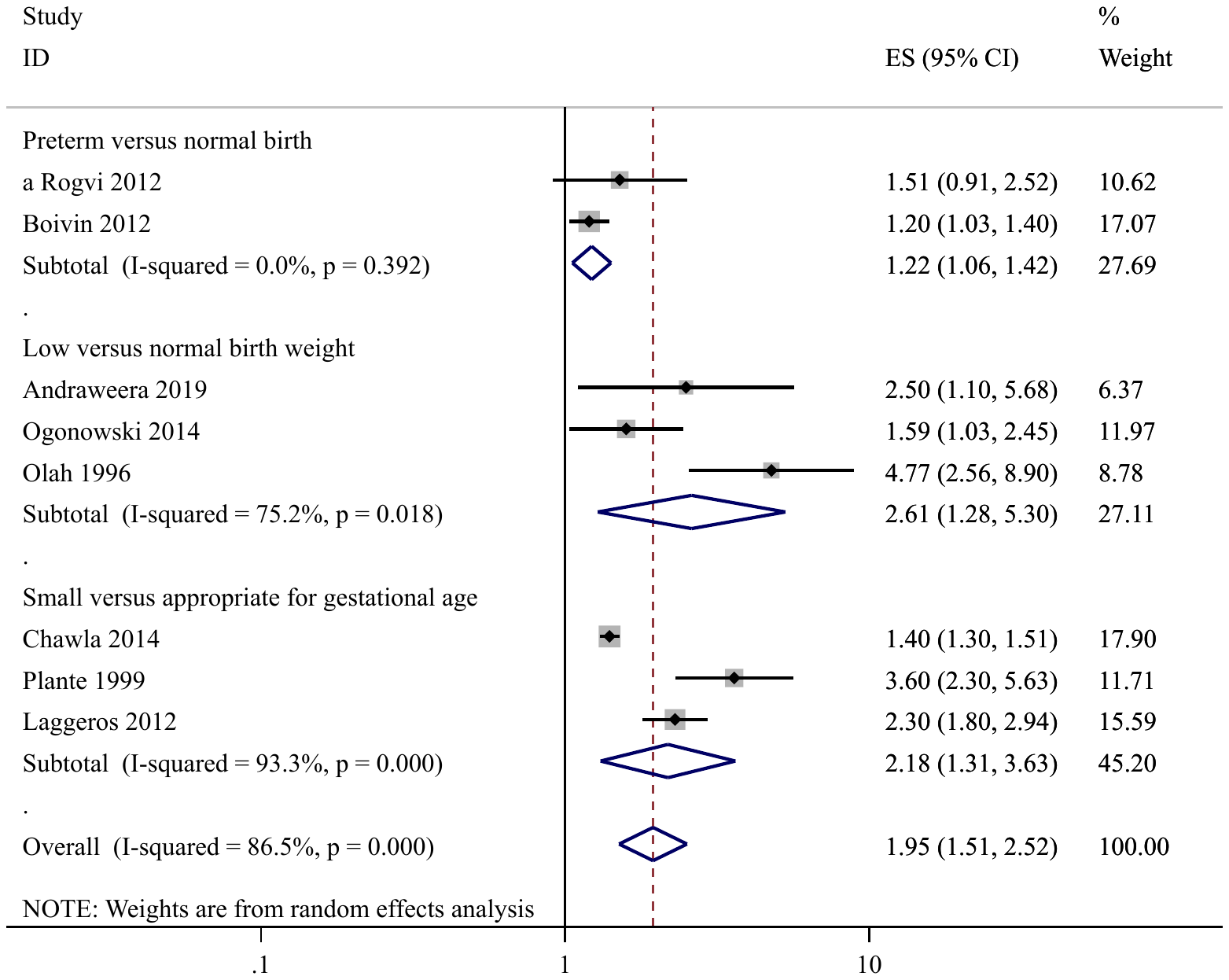

**Supplementary Figure S5**

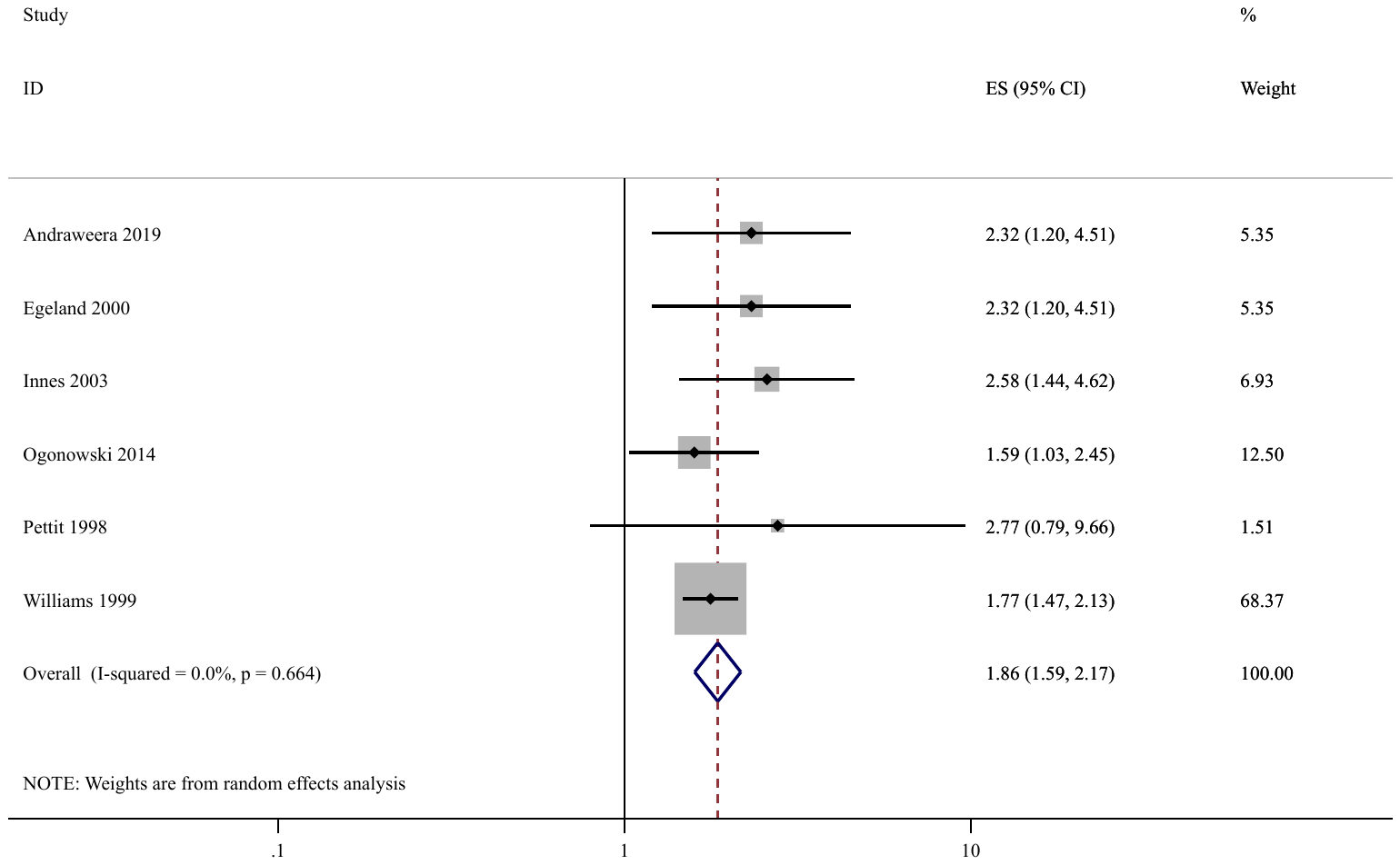
